# Chemokine Receptor 2 Is A Theranostic Biomarker for Abdominal Aortic Aneurysms

**DOI:** 10.1101/2023.11.06.23298031

**Authors:** Santiago Elizondo-Benedetto, Sergio Sastriques-Dunlop, Lisa Detering, Batool Arif, Gyu Seong Heo, Deborah Sultan, Hannah Luehmann, Xiaohui Zhang, Xuefeng Gao, Kitty Harrison, Dakkota Thies, Laura McDonald, Christophe Combadière, Chieh-Yu Lin, Yeona Kang, Jie Zheng, Joseph Ippolito, Richard Laforest, Robert J. Gropler, Sean J. English, Mohamed A. Zayed, Yongjian Liu

## Abstract

Abdominal aortic aneurysm (AAA) is a degenerative vascular disease impacting aging populations with a high mortality upon rupture. There are no effective medical therapies to prevent AAA expansion and rupture. We previously demonstrated the role of the monocyte chemoattractant protein-1 (MCP-1) / C-C chemokine receptor type 2 (CCR2) axis in rodent AAA pathogenesis via positron emission tomography/computed tomography (PET/CT) using CCR2 targeted radiotracer ^64^Cu-DOTA-ECL1i. We have since translated this radiotracer into patients with AAA. CCR2 PET showed intense radiotracer uptake along the AAA wall in patients while little signal was observed in healthy volunteers. AAA tissues collected from individuals scanned with ^64^Cu-DOTA-ECL1i and underwent open-repair later demonstrated more abundant CCR2+ cells compared to non-diseased aortas. We then used a CCR2 inhibitor (CCR2i) as targeted therapy in our established male and female rat AAA rupture models. We observed that CCR2i completely prevented AAA rupture in male rats and significantly decreased rupture rate in female AAA rats. PET/CT revealed substantial reduction of ^64^Cu-DOTA-ECL1i uptake following CCR2i treatment in both rat models. Characterization of AAA tissues demonstrated decreased expression of CCR2+ cells and improved histopathological features. Taken together, our results indicate the potential of CCR2 as a theranostic biomarker for AAA management.

**Graphical Abstract:** 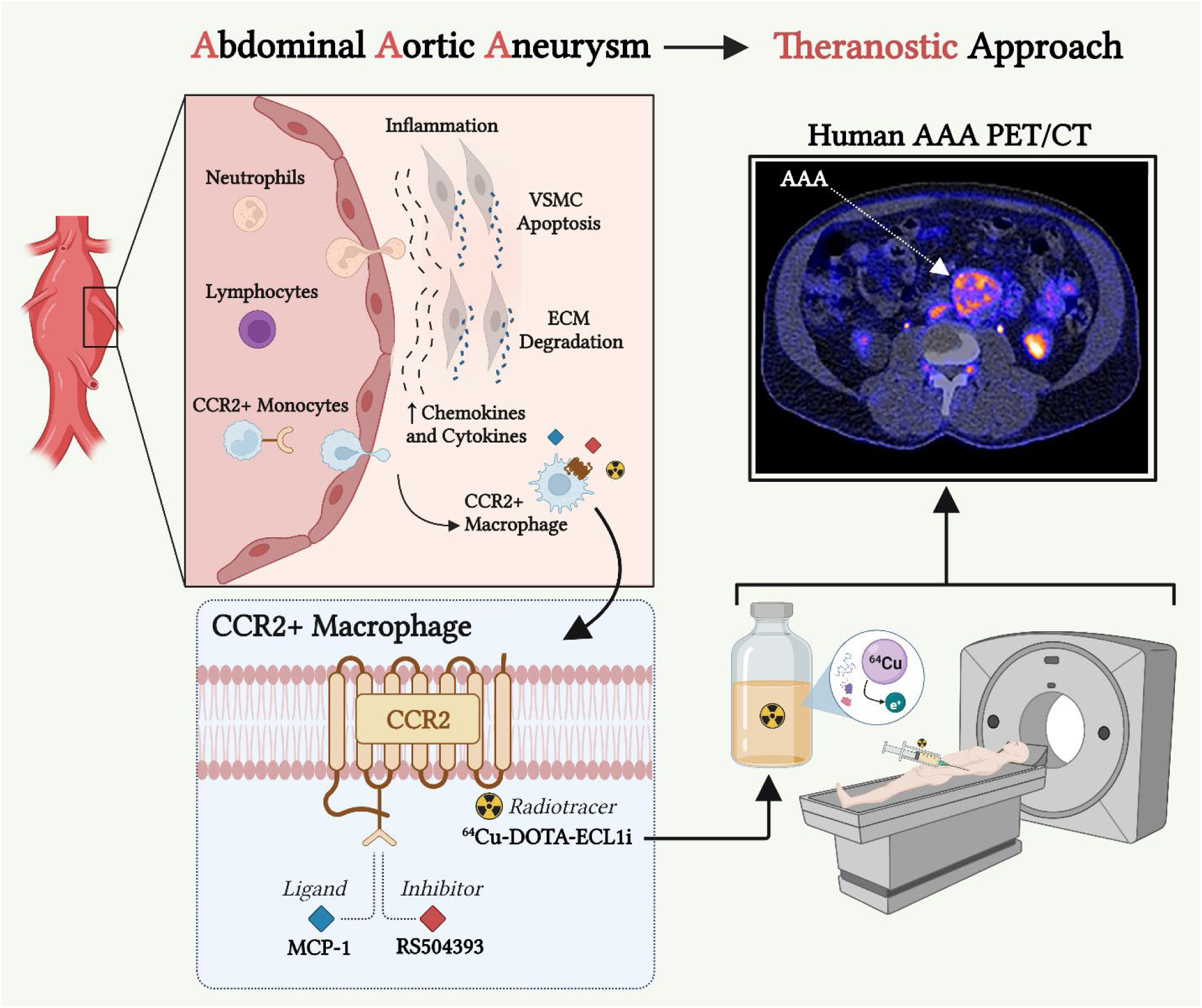

## Introduction

Abdominal aortic aneurysms (AAA) represent a potentially life-threatening condition as they typically remain asymptomatic until they subsequently rupture. This unpredictable outcome can lead to a high morbidity and mortality rate, and substantial health care costs (1, 2). AAAs are more prevalent in men, but they rupture more often in women for unknown reasons (3, 4). Although risk factors such as hypertension and smoking are linked to AAA formation, the underlying mechanisms for expansion and risk of rupture remains a topic of intense investigation (5, 6).

Clinical screening and surveillance protocols for AAAs continue to be significantly limited as they are almost entirely based on anatomic diameter measurements that do not account for the complex molecular processes occurring within AAA tissue (3, 7). Additionally, there remains no effective medical therapies that can impede AAA expansion and the risk of rupture (3, 8-10). Invasive surgical management continues to be the only option for individuals who meet the traditional size criteria for AAA repair or who develop AAA-related symptoms (11). Therefore, current AAA management leaves patients in prolonged anticipation of either an impending need for a future repair, or risk of rapid progressing to rupture which can be life-threatening (12-14).

Various studies have identified inflammation as a key feature associated with AAA pathogenesis (15-18). The presumed process is that cytokines and chemokines are secreted by immune cells following vascular injury, which in turn promotes recruitment of additional inflammatory cells, such as monocytes and macrophages, and further infiltration in the aortic tissues that cause further damage. Increased tissue oxidative stress and local release of matrix metalloproteinases (MMPs) is thought to trigger vascular smooth muscle cells (VSMCs) apoptosis and extracellular matrix (ECM) degradation, which results in AAA expansion (19-21). During this process, structural proteins such as elastin and collagen lose their integrity, causing weakness within the aortic wall architecture, further promoting the inflammatory cascade and AAA sac expansion (18, 19). As such, recent studies have investigated whether targeting inflammation signaling pathways can decrease AAA expansion and prevent its rupture (22-26). Despite early promising data in rodent models, to date there has been very limited translation in humans with AAA, and therefore the search for an inflammation-mediating strategy for AAA management remains critically needed (27).

The search for specific biomarkers to enable molecular imaging and targeted therapy of AAA has been an on-going effort to address this unmet clinical need. Many imaging agents have been developed for AAA detection in preclinical and/or clinical research showing promising results such as matrix metalloproteinase (MMP) and integrin (28-30). Moreover, a theranostic strategy that allows for both real-time detection of vulnerable AAA as well as applying targeted therapeutics to blunt disease progression has never been realized. Our previous work suggests that targeting chemokines and their cognate receptors that regulate and guide leukocytes migration to the site of vascular injury may be a feasible strategy (31). Of various chemokine receptors, chemokine receptor type 2 (CCR2) and its natural ligand, monocyte chemoattractant protein-1 (MCP-1), play a central role in cell signaling that promotes leukocytes trafficking to the arterial wall during the initiation and progression of AAA (32-34). Pharmacological intervention using CCR2 inhibitor (CCR2i) or genetic depletion of CCR2 could significantly decrease the quantity of pro-inflammatory monocytes and attenuate the dilatation and rupture of aneurysms in AAA mice (35, 36), underlining the role of CCR2 in AAA treatment.

We recently demonstrated that CCR2+ monocytes/macrophages were highly elevated in the aortic wall of human AAA tissues. In a ruptured male AAA (MRAAA) rat model, our novel CCR2 targeted radiotracer, ^64^Cu-DOTA-ECL1i, demonstrated the accurate detection of AAA and the potential to predict the rupture of AAA using positron emission tomography/computed tomography (PET/CT), highlighting the capability of CCR2 as a theranostic biomarker for AAA management (37).

In the current study, we hypothesize that CCR2 may serve as a suitable theranostic biomarker for AAA tissue. We successfully translated ^64^Cu-DOTA-ECL1i into humans and performed proof-of-concept PET/CT studies in AAA patients and healthy volunteers (HV). These studies were complemented with assessment of ^64^Cu-DOTA-ECL1i imaging in AAA rat models and evaluation of the therapeutic effect of CCR2 antagonism on AAA expansion and rupture. In contrast to the low radiotracer retention in HVs, ^64^Cu-DOTA-ECL1i revealed intense and heterogeneous tracer uptake within the aneurysm of AAA patients. The aneurysmal tissues collected from imaged AAA patients underwent open repair a few weeks later showed abundant expression of CCR2+ cells. To explore the effect of sex differences in AAA imaging and therapy, we next established a novel ruptured female AAA rat model (FRAAA). PET/CT with ^64^Cu-DOTA-ECL1i revealed sensitive and specific detection of CCR2 within FRAAA and demonstrated its potential to predict the rupture of FRAAA. Moreover, we used a highly selective CCR2 antagonist (RS504393) for the pharmacological intervention of AAA rupture in both male and female rat models. We demonstrated the effectiveness of this strategy for preventing AAA rupture and improvement of the histopathological features of AAA (38), which was consistent with the reduction of CCR2 PET signals.

## Results

### First-in-human ^64^Cu-DOTA-ECL1i PET/CT showed increased tracer uptake in AAA tissue

CCR2 PET/CT was carried out in HVs and patients with AAAs (Figure 1, Supplemental Figure 1, and Table 1). Consistent with our previous findings (39), ^64^Cu-DOTA-ECL1i demonstrated fast renal and blood clearance and low retention in major organs. In contrast to the minimal ^64^Cu-DOTA-ECL1i retention in the infrarenal aorta of HV subjects, the CCR2 radiotracer revealed intense and heterogeneous uptake at the intraluminal surface of AAA, particularly at regions mostly adjacent to calcified atherosclerotic plaque within the AAA wall (Figure 1A, Supplemental Figure 1). Minimal PET signals were determined in the AAA sac thrombus as identified on computed tomography angiography (CTA) images. Quantitative uptake analysis showed both the maximal and mean standardized uptake values (SUVmax and SUVmean) in patients with AAA were more than twice those determined in HVs (Figure 1B), supporting ^64^Cu-DOTA-ECL1i imaging specificity in AAA tissue. Moreover, we performed a kinetic analysis of ^64^Cu-DOTA-ECL1i uptake in AAA patients and HV subjects and determined the distribution volume (DV) using logan plot. As shown in Supplemental Figure 2, the DV values of AAA patients, calculated by evaluating multiple image-derived input functions from either the aorta or vena cava, were significantly higher than those determined from HV subjects, further supporting ^64^Cu-DOTA-ECL1i targeting specificity (Figure 1A).

**Figure 1.**
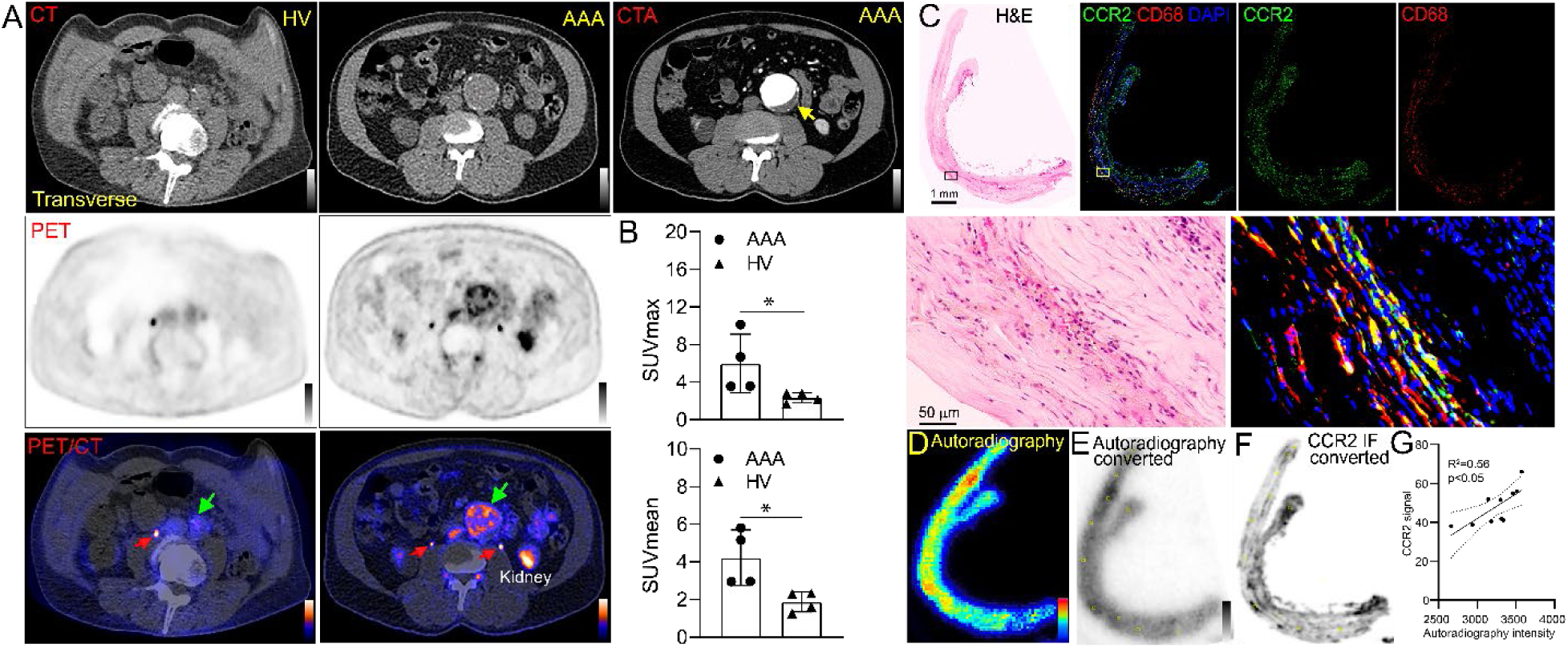
^64^Cu-DOTA-ECL1i PET/CT images in a healthy volunteer (HV) and a patient with AAA, and histopathological characterization of aneurysmal tissue. (**A**) Transverse view of CT, PET and PET/CT fused images of the infrarenal aorta of a HV (male) and an AAA patient (male) both at their 60s with additional CTA image. In contrast to the low tracer retention in the infrarenal aorta of HV, ^64^Cu-DOTA-ECL1i showed strong uptake along the intraluminal surface of the aneurysm. Red arrow: ureter; Green arrow: AAA; Yellow: thrombus. (**B**) Quantitative uptake analysis of ^64^Cu-DOTA-ECL1i in infrarenal artery of the HV and AAA of the patient. (**C**) Whole tissue and Zoomed-in images of H&E and IF staining of aneurysmal tissue removed from the AAA patient during open repair. H&E showed significant amount of infiltrated inflammatory cells. Whole tissue and zoomed-in IF staining images of the AAA tissue demonstrated abundant expression of CCR2+ (green) cells that largely co-localized with CD68 (red). (**D**) Autoradiography of ^64^Cu-DOTA-ECL1i binding to human AAA tissue. Converted low-resolution images of autoradiography (**E**) and immunofluorescence staining of CCR2 (**F**). The two sets of images were co-registered with representative regions-of-interest (ROIs, yellow squares on E and F). (**G**) Signals extracted from the ROIs of the two sets of images showed linear correlation. Data presented as Mean ± SD. *p < 0.05, **p < 0.01, ***p < 0.001, **** p<0.0001.

**Table 1.** Demographics and ^64^Cu-DOTA-ECL1i uptake in healthy volunteers and patients with AAA*.

| **Subject** | **Age**<br>**(year)** | **Sex** | **Surgery** | **AAA size**<br>**(cm)** | **Hypertension** | **Tobacco** | **SUV<sub>max</sub>** | **SUV<sub>mean</sub>** |
| --- | --- | --- | --- | --- | --- | --- | --- | --- |
| **HV-1** | 70s | F | N/A | N/A | Y | Former | 2.82 | 2.34 |
| **HV-2** | 70s | F | N/A | N/A | Y | N | 2.79 | 2.30 |
| **HV-3** | 60s | M | N/A | N/A | N | N | 2.15 | 1.61 |
| **HV-4** | 30s | F | N/A | N/A | N/A | Former | 1.70 | 1.26 |
| **AAA-1** | 60s | M | Open repair | 5.5 | Y | N | 10.1 | 5.81 |
| **AAA-2** | 50s | M | Open repair | 9.0 | Y | Current | 3.56 | 2.95 |
| **AAA-3** | 60s | M | Open repair | 5.5 | Y | N | 6.69 | 5.16 |
| **AAA-4** | 70s | F | Open repair | 5.4 | Y | Former | 3.58 | 2.97 |

\*All subjects were non-diabetic.

The four AAA patients enrolled in this study had aortic maximum diameters ranging from 5.4 cm to 9.0 cm and were all hypertensive but not diabetic (Table 1). However, tracer uptake showed little correlation with the AAA diameters and were independent of the consumption of tobacco. Histopathological analysis of AAA tissue collected from patients who underwent ^64^Cu-DOTA-ECL1i PET/CT scan and then open AAA repair revealed significant infiltration of inflammatory cells. Whole tissue immunofluorescence (IF) staining and magnified images showed high expression of CCR2+ cells throughout the tissue, which were also largely positive for CD68, consistent with our previous report (Figures 1C) (37). *Ex vivo* autoradiography revealed intense radiotracer binding to the AAA tissue in a pattern similar to CCR2 expression determined by IF staining (Figure 1D). Regions-of-interest (ROIs) based analysis of converted autoradiography (Figure 1E) and CCR2 IF image (Figure 1F) demonstrated a significant linear correlation, indicating the binding specificity of ^64^Cu-DOTA-ECL1i to human CCR2 (Figure 1G) (39). *Characterization of CCR2 in human AAA ruptured tissues.* To further assess the role of CCR2 in AAA rupture, we compared three retrospective groups of biobanked human tissue samples including normal abdominal aorta (NAA), AAA, and ruptured AAA (RAAA). The NAA showed an intact elastic laminae and a minimal amount of inflammatory cells in the tunica intima and tunica media. Consistent with the histology finding, IF staining revealed few CCR2+ or CD68+ cells (Supplemental Figure 3A). Both AAA and RAAA specimens demonstrated hemorrhage, significant loss of VSMCs, disruption of elastic laminae, increased collagen deposition in the tunica media, substantial infiltration of inflammatory cells, and formation of luminal thrombus (Supplemental Figure 3B & C). IF staining indicated extensive and heterogeneous expression of CCR2+ cells throughout both AAA and RAAA specimens with more CCR2+ and CD68+ cells observed in RAAA tissues, especially in the regions with atherosclerosis (Supplemental Figure 3D). *Ex vivo* autoradiography and correlation analysis also demonstrated the binding specificity of ^64^Cu-DOTA-ECL1i to the RAAA tissue (Supplemental Figure 3E-H).

### ^64^Cu-DOTA-ECL1i PET shows potential to predict AAA rupture in a female AAA rat model

In humans, AAA rupture happens more frequently in females than males (4) Based on our previous studies about CCR2 in male AAA rats (37), we developed a female rat AAA model to evaluate the impact of CCR2 on AAA formation, expansion, and rupture In wild type non-operated female rats (WT), ^64^Cu-DOTA-ECL1i PET/CT displayed rapid renal clearance and low retention in the abdominal aorta (AA). At day 7 post surgery, low tracer uptake was determined in the AA of sham-operated female rats (sham) while an intense signal was localized in AAA-induced female rats (FAAA) (Figure 2A). Interestingly, increased tracer uptake in FAAA rats was also observed at 14 days post injury (Figure 2B), suggesting persistent expression of CCR2 in the FAAA rat model. Compared to the tracer uptake values determined in male wild type, sham and MAAA rats (37), ^64^Cu-DOTA-ECL1i uptake in the corresponding female rat models displayed a similar pattern, highlighting the validity of our FAAA models (Figure 2C). Moreover, *ex vivo* autoradiography of FAAA rat tissue demonstrated strong and heterogeneous tracer uptake within the aneurysm but minimal accumulation in the thoracic aorta and psoas muscle, suggesting the specific binding of ^64^Cu-DOTA-ECL1i to FAAA. In sham operated rats, low uptake of ^64^Cu-DOTA-ECL1i was observed in the abdominal aorta (Figure 2D).

**Figure 2.**
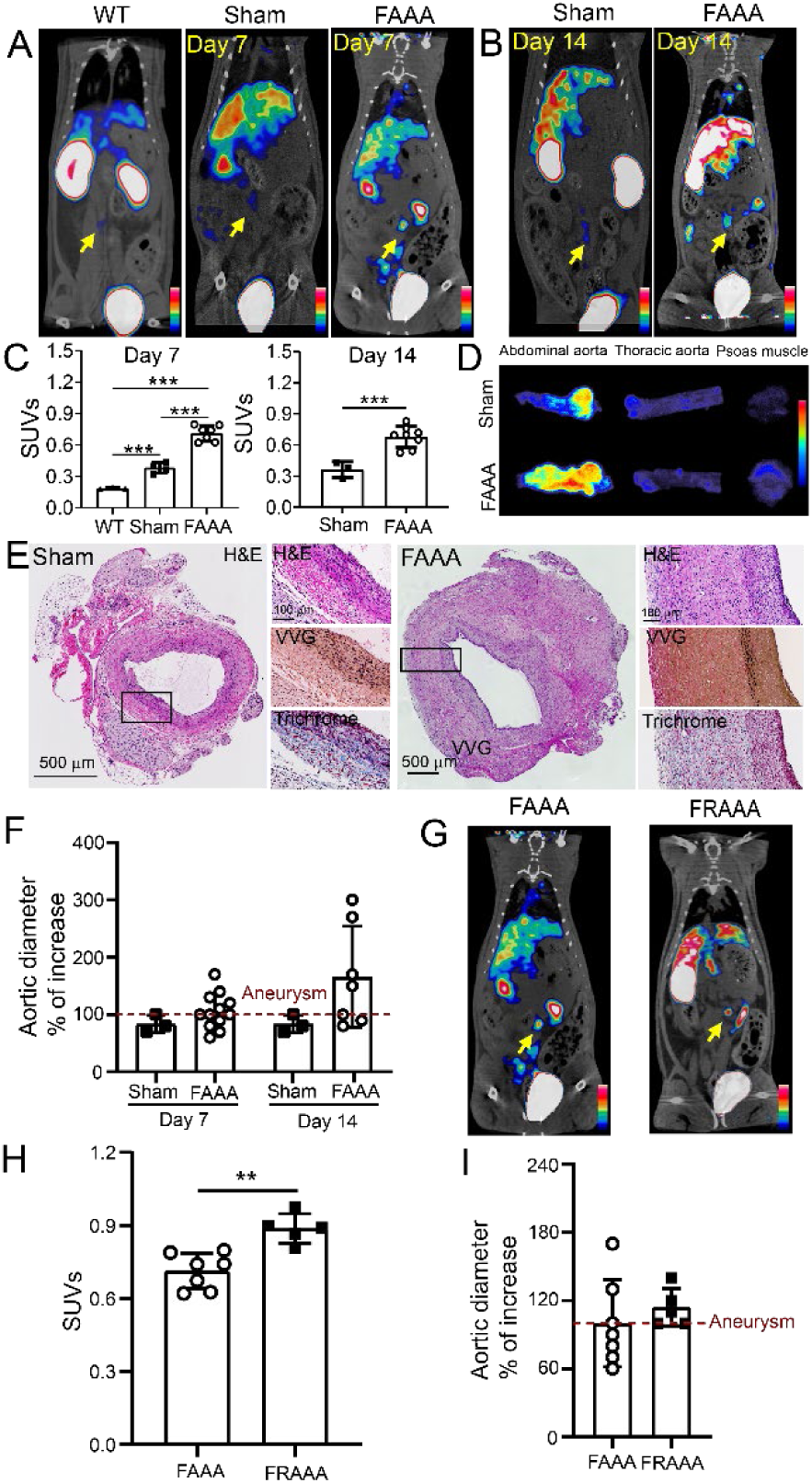
Assessment of CCR2 PET imaging AAA and rupture potential in a female rat model. Representative ^64^Cu-DOTA-ECL1i PET/CT images for (**A**) WT, sham-control, and FAAA female rats at day 6 post PPE exposure and (**B**) Sham-control and FAAA female rats at day 14 post PPE exposure with an intense signal detected within the aneurysm of the FAAA rats (yellow arrow). (**C**) Quantitative tracer uptake of ^64^Cu-DOTA-ECL1i at day 6 and day 14 in WT rats (n=3), sham-controls (n=3-4) and FAAA (n=7-8) respectively. (**D**) *Ex vivo* autoradiography of ^64^Cu-DOTA-ECL1i in thoracic aortas, abdominal aortas, and psoas muscle of FAAA and sham-control rats. (**E**) H&E, VVG and MT trichrome staining of abdominal aortas (cross-section of tissue slides) with 5 × magnification and 10 × magnification in sham and FAAA groups respectively. (**F**) Percent increases of aortic diameters determined by ultrasound showing increased sizes in FAAA rats than those in sham-controls. Red dotted line is set at 100%. (**G**) Representative ^64^Cu-DOTA-ECL1i PET/CT images at day 6 post PPE exposure showed significant signal in FRAAA rats (yellow arrow) compared to non-ruptured FAAA rats. (**H**) Quantification of tracer uptake in FRAAA rats showing significantly higher signals than that in FAAA rats at day 6 post PPE exposure. (**I**) Percent increases of aortic diameters determined by ultrasound showing comparable sizes between FAAA and FRAAA rats at day 6 post PPE. Data presented as Mean ± SD. *p < 0.05, **p < 0.01, ***p < 0.001, **** p<0.0001.

When compared to sham operated female rats, histopathological assessment by H&E, Verhoeff-Van Gieson (VVG), and trichrome stains showed that aneurysmal tissue collected from FAAA rats at day 14 had dilated aortic diameter, significant loss of VSMCs, disrupted elastic lamina, increased infiltration of inflammatory cells, and significant adventitial fibrosis. These histopathologic features of FAAA rats largely recapitulated findings observed in human AAAs, lending support to use of the β-aminopropionitrile (BAPN) rat rupture model as a useful platform for imaging human AAA associated inflammation (Figure 2E). Moreover, sham-operation induced enlargement of the aortic diameter in female rats was stable from days 7-14 as measured by ultrasound. In contrast, the sizes of FAAAs continuously increased during the 14-day study (Figure 2F). Taken together, the AAA histopathological features, variation of aortic diameter following injury, and ^64^Cu-DOTA-ECL1i PET imaging demonstrate the feasibility of the FAAA rat model for theranostic applications.

Previously, we reported the potential of CCR2 PET imaging AAA rupture in MRAAA rats (37). To further validate the potential of ^64^Cu-DOTA-ECL1i imaging AAA vulnerability, we established a female raptured AAA (FRAAA) rat model following the similar surgical procedures used for MRAAA rats. Interestingly, FRAAA rats showed a comparable AAA rupture kinetics to MRAAA, with rupture occurring between days 6 and 10 post surgery. ^64^Cu-DOTA-ECL1i PET/CT revealed higher tracer uptake within the FRAAA than those in the non-ruptured FAAA rats, while the increase in aortic diameter was comparable between the two groups (Figure 2G, H, I), further confirming the potential of CCR2 PET for predicting the rupture in the FRAAA rat model.

### CCR2 inhibition attenuates AAA rupture and can be visualized by ^64^Cu-DOTA-ECL1i PET/CT

Due to the known role of the MCP-1/CCR2 axis in mediating leukocyte trafficking, CCR2 antagonism has been widely used as a pharmacological intervention in both preclinical and clinical research for oncology, neuroinflammation, and cardiovascular diseases (40-44). We therefore evaluated whether CCR2 antagonism with RS504393 (45, 46), a highly selective CCR2 inhibitor (CCR2i), could modulate the expansion and risk of AAA rupture in both MRAAA and FRAAA rat models. The initial treatment was carried out in MRAAA rats using RS504393 administered via oral gavage prior to MRAAA induction (CCR2i D0) and compared to those without RS504393 treatment (Figure 3A). In contrast to an incidence of 66% rupture rate observed in MRAAA rats at day 14, the rupture rate in the CCR2i D0 group was significantly decreased with significantly improved survival (Supplemental Figure 4A & B). Moreover, CCR2i treatment substantially inhibited the expansion of MRAAA diameter by approximately 100% relative to the non-treated group at day 7, which was further inhibited at day 14 (Supplemental Figure 4C). Importantly, ^64^Cu-DOTA-ECL1i PET/CT performed at day 7 and 14 in CCR2i D0 rats revealed significantly reduced tracer uptake compared to MRAAA rats without treatment (Supplemental Figure 4D & E), which was consistent with decreased AAA sizes and improved survival.

**Figure 3.**
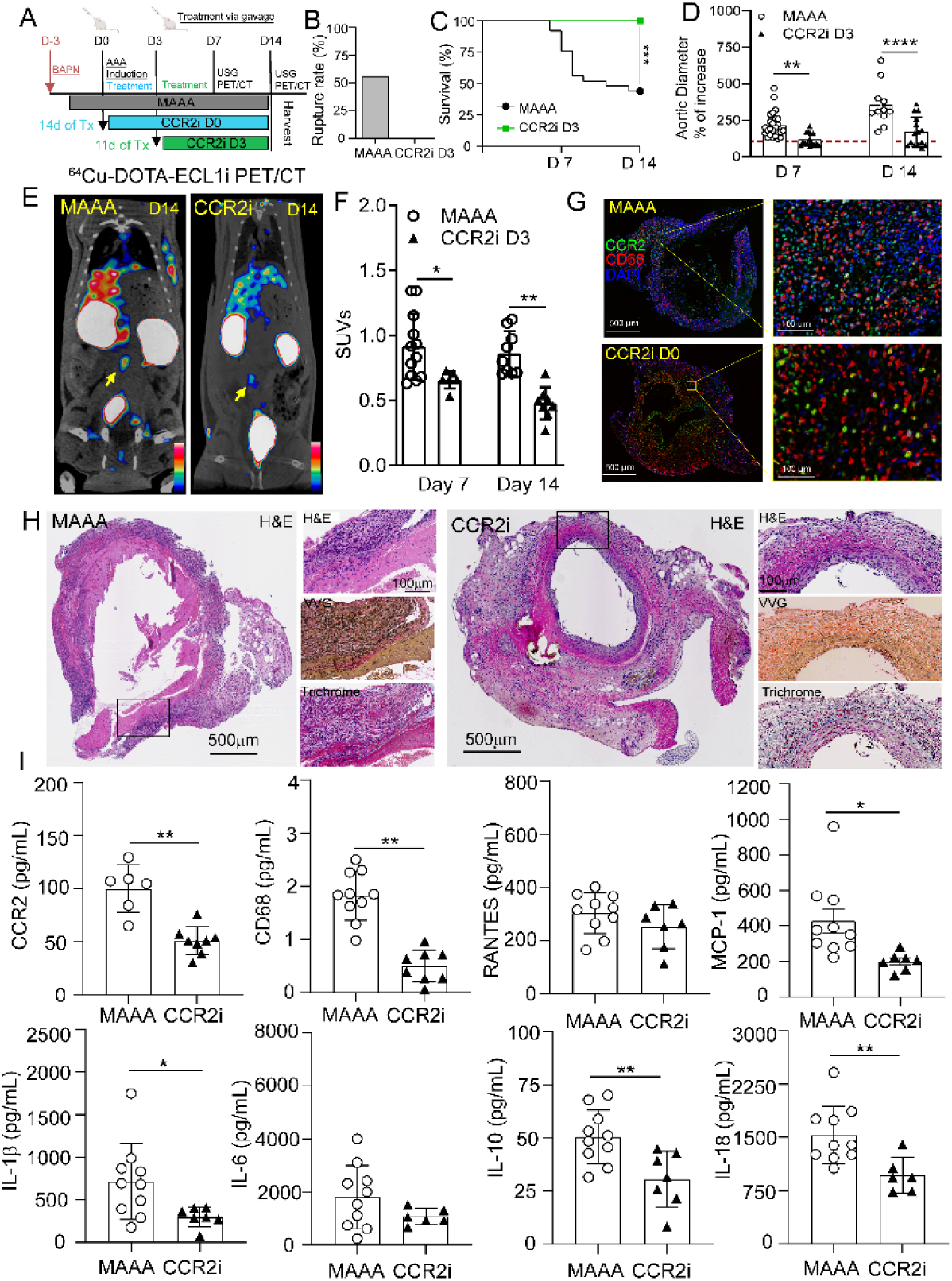
Assessment of CCR2 inhibition on AAA progression and rupture in a MRAAA rat model. (**A**) Diagram showing the MRAAA (n=25) rat model and treatment strategies using CCR2 inhibitor RS504393 (CCR2i) starting at day 0 (n=19) and day 3 (n=15) post elastase exposure. (**B**) Comparison of AAA rupture rates between MRAAA rats without treatment and those treated with CCR2i starting at day 3 (CCR2i D3). (**C**) Kaplan-Meier curve demonstrating significantly improved survival of MRAAA rats following CCR2i D3 treatment. (**D**) Percent aortic diameter increase in MAAA vs CCR2i D3 groups at day 7 (207 ± 71 vs 117 ± 44) and day 14 (297 ± 74 vs 171 ± 102), respectively. Red dotted line is set at 100%. (**E**) Representative ^64^Cu-DOTA-ECL1i PET/CT images at day 14 post PPE exposure showed specific and intense detection of AAA (yellow arrow) in MAAA, compared with the low trace accumulation in the CCR2i D3 treated group. (**F**) Quantitative tracer uptake in MAAA rats without and with CCR2i D3 treatment at day 7 (MAAA: n=12, CCR2i D3: n=6) and day 14 (MAAA: n=9, CCR2i D3: n=8) post PPE. (**G**) IF and (**H**) H&E, trichrome and VVG staining of AAAs (cross-sectional) collected from MAAA rats and those following CCR2i D3 treatment. (**I)** ELISA assays of inflammatory cytokines in MAAA rats with and without CCR2i D3 treatment. Data presented as Mean ± SD. *p < 0.05, **p < 0.01, ***p < 0.001, **** p<0.0001.

We next assessed the time-dependence of RS504393 treatment in MRAAA rats by evaluating the impact of CCR2 inhibition starting from day 3 (CCR2i D3) post induction of AAA. Notably, all CCR2i D3 rats survived during the 14-day study and demonstrated a 0% rupture rate (Figure 3B & C) and the aortic diameters of CCR2i D3 rats were significantly lower than those in MAAA group (Figure 3D). Moreover, quantitative uptake analysis of CCR2 PET/CT images revealed significantly decreased tracer accumulation within the AAAs of CCR2i D3 rats compared to the non-treated MAAA group (Figure 3E & F), indicating the potential of ^64^Cu-DOTA-ECL1i imaging to assess AAA vulnerability, prospective rupture, and treatment outcome.

IF staining and enzyme-linked immunosorbent assay (ELISA) were performed to characterize the variation of CCR2 expression and inflammatory cytokine abundance, important factors in AAA development following CCR2i treatment (47, 48). As shown in Figure 3G and Supplemental Figure 5A, quantification of IF images showed that inhibition of CCR2 led to significantly decreased CCR2+ cells in AAA tissue. In contrast to the histopathological features observed in MAAA rats, including significant disruption of the elastic lamina, deposition of collagen in the tunica intima and media, loss of VSMCs, and notable infiltration of inflammatory cells, we observed less elastin degradation, minimal loss of VSMCs, and reduced fibrosis in the CCR2i D3 rats (Figure 3H, Supplemental Figure 6A&B, and Supplemental Figure 7). Moreover, ELISA assays showed that RS504393 inhibition led to approximately 50% reduction of CCR2 and 70% of CD68 content in CCR2i D3 rats compared to the non-treated MAAA rats at day 14 (Figure 3I). Additionally, MCP-1, IL-10, and IL-18 expressions were all significantly decreased following CCR2i treatment, despite RANTES and IL-6 levels being unchanged between the two groups (Figure 3I). These findings were consistent with the reported anti-inflammatory properties of RS504393 (41, 45, 46).

### CCR2 inhibition significantly attenuates AAA rupture in female RAAA rats

Clinically, the management of female AAA patients is more challenging since their AAAs usually present late and grow more rapidly, resulting in a four-fold increase in risk of rupture and worse outcomes than their male counterparts (4, 49). Based on the effective prevention of AAA rupture in male rats using RS504393, we evaluated whether CCR2i starting from day 3 post induction could influence AAA growth and rupture in FRAAA rats. Consistent with our findings in MRAAA rats, RS504393 demonstrated effective prevention of rupture and significantly improved survival at day 14 in FRAAA rats (Figure 4 A & B). Furthermore, the aortic diameters in CCR2i D3 FRAAA rats were significantly lower than those in FAAA rats, similar to their male counterparts (Figure 4C). Importantly, ^64^Cu-DOTA-ECL1i PET/CT uptake of CCR2i D3 FRAAA group was decreased approximately 50% relative to that in non-treated FAAA rats (Figure 4 D& E), which was consistent with the data collected in MRAAA CCR2i D3 rats.

**Figure 4.**
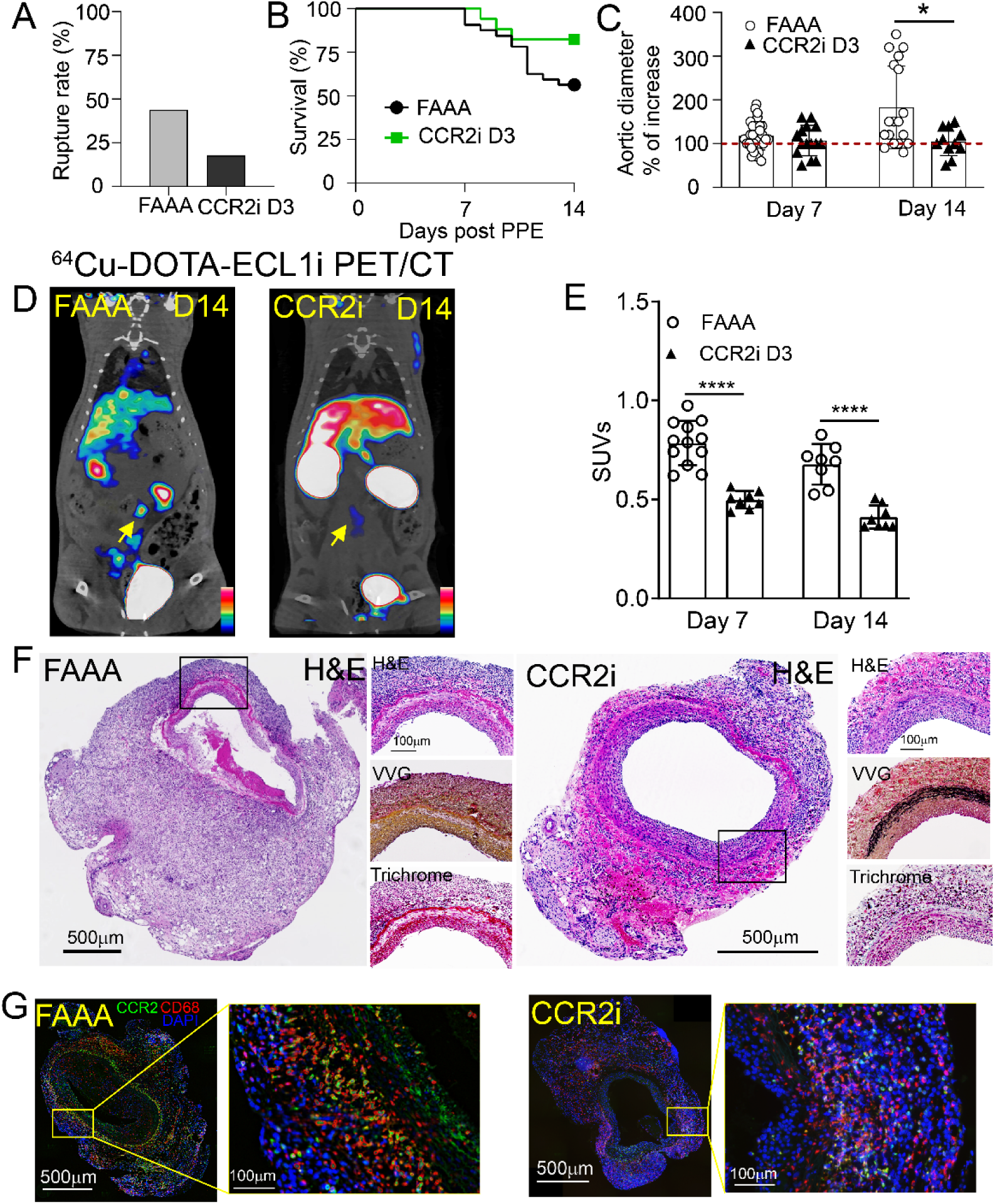
Assessment of CCR2 inhibition on AAA progression and rupture in a *F*RAAA rat model. **(A)** AAA rupture rate*s* in FAAA (n=32) and CCR2i D3 (n=17) groups. (**B**) Kaplan-Meier curve demonstrating improved survival following CCR2i D3 treatment in FRAAA rats. (**C**) Percent aortic diameter increase in FAAA vs CCR2i D3 treated rats at day 7 (117 ± 32 vs 106 ± 35) and day 14 (183 ± 94 vs 104 ± 32, p=0.01). Red dotted line is set at 100%. (**D**) Representative ^64^Cu-DOTA-ECL1i PET/CT images at day 6 post PPE exposure showed specific and intense detection of AAA (yellow arrow) in FAAA while low tracer accumulation was observed in the CCR2i D3 treated group. (**E**) Quantitative tracer uptake in FAAA rats without and with CCR2i D3 treatment at day 7 (FAAA: n=12, CCR2i D3: n=8) and day 14 (FAAA: n=8, CCR2i D3: n=8) post PPE. (**F**) H&E, trichrome and VVG staining and IF (G) of AAAs (cross-sectional) collected from *F*AAA rats and those following CCR2i D3 treatment. Data presented as Mean ± SD. *p < 0.05, **p < 0.01, ***p < 0.001, **** p<0.0001.

Histopathological analysis of AAAs in FRAAA CCR2i D3 rats demonstrated relatively preserved elastic laminae, minimal loss of VSMCs, and compensatory medial hypertrophy compared to observations in untreated FAAA rats (Figure 4F). IF staining revealed significantly reduced CCR2+ cells and a modest decrease of CD68+ macrophages (Figure 4G, Supplemental Figure 6C&D & Supplemental Figure 7), consistent with the ^64^Cu-DOTA-ECL1i PET/CT findings (Figure 4 D& E).

## Discussion

Despite remarkable progress in the surgical management of AAAs, effective pharmacological treatment of individuals who do not yet meet the size criteria for operative repair are still lacking. This inability to retard AAA growth and prevent rupture in individuals who are receiving expectant management continues to be a significant unmet clinical need. A theranostic biomarker that can be used for both diagnosis and therapy can be of significant clinical value to address these challenges. Recent studies suggest that AAA pathophysiology is a multifactorial process initiated by inflammatory stimuli that leads to a cascade of actions including MMP activation, oxidative stress, intraluminal thrombus formation, VSMCs apoptosis, and ECM degeneration (16, 50-53). Therefore, much effort has been dedicated to target inflammatory cells as a potential strategy to uncover effective diagnostic and therapeutic strategies to improve the management of AAAs (25, 50, 53-55).

Of the various biomarkers expressed by monocytes/macrophages, the MCP-1/CCR2 axis plays an important role in monocyte recruitment, activation, macrophage differentiation, promoting inflammation in vascular tissue, and ECM remodeling, leading to AAA instability. Previously, we demonstrated the sensitivity and specificity of ^64^Cu-DOTA-ECL1i PET/CT for detecting unstable AAA and predicting its rupture in a MRAAA rat model (37). Others have shown that CCR2 inhibition decreased the infiltration of macrophages, secretion of inflammatory cytokine, and the formation of AAAs in murine AAAs (34, 36). Collectively these findings highlighted the potential of CCR2 as a theranostic biomarker for AAA diagnosis and therapy.

The management of female patients with AAAs is notably more challenging than their male counterparts. AAAs in female patients are typically smaller but have a higher rate of rupture (4). Preclinical rodent studies demonstrated that differences in AAA stability may be associated with female sex hormones, but the molecular mechanisms of action are still elusive (56). So far, several rodent female AAA rupture models have been established that demonstrated the pathological and clinical features mimicking female AAA patients. However, their rupture rates were low and the rupture kinetics were significantly delayed compared to the corresponding male AAA rupture models (56-58), which is opposite to what happens in humans. Based on our previous studies (37), here we report a novel elastase-induced FRAAA rat model showing location-specific infrarenal AAA and a consistent rate of rupture. Similar to the well-established MRAAA rat model, the rupture rate and kinetics of the as-developed FRAAA rat model were comparable, which allowed for a unique opportunity to comparatively study biomarkers that can impact AAA disease progression in both male and female counterparts. Histopathological characterization of FRAAA tissues revealed typical features of AAA including adventitial thickening, fibrosis, leukocyte infiltration, VSMCs apoptosis, luminal expansion, and ECM degradation (57).

An unmet clinical challenge for the management of AAA patients is to sensitively and specifically determine AAA progression and assess its vulnerability in a non-invasive manner. Clinical imaging modalities for AAAs such as ultrasound, CT, and magnetic resonance imaging (MRI) mainly focus on the anatomic features (diameter, volume) of the AAA. Although new imaging parameters such as AAA volume and aortic size index (ratio of aortic diameter and body surface area) are being explored, their applications to predict the risk of AAA rupture are currently limited (59-61). Recently, a multi-center study using ultrasmall superparamagnetic particles of iron oxide-enhanced MRI demonstrated potential to predict the rate of AAA growth and clinical outcome. However, the assessment needs to be integrated with established clinical factors (62). Functional imaging-based modalities such as ^18^F-FDG and ^18^F-sodium fluoride have demonstrated some promise in patients with AAAs (29, 63) but lack both specificity for critical determinants of the inflammatory response in AAA development and progression and theranostic potential for patients undergoing expectant management.

In contrast to anatomy-based imaging modalities, molecular imaging affords the potential to non-invasively track cell surface biomarkers to gain insight into AAA pathogenesis and to uncover underlying mechanisms. Although some molecular agents have shown promising results for AAA imaging in male murine models, their application in female AAA models has not yet been tested (28, 29). Compared to the low background retention of ^64^Cu-DOTA-ECL1i in the descending aortas of wild type and sham-operated rats, CCR2 PET/CT in FAAA rats showed an intense and specific uptake within the AAA at both time points evaluated, which was further confirmed by *ex vivo* autoradiography. Moreover, in the ruptured FRAAA rats, ^64^Cu-DOTA-ECL1i revealed a significantly higher accumulation within the aneurysm than the non-ruptured FAAA, which was independent of the aortic diameters. These results were consistent with what we previously reported in MRAAA rats, highlighting the uniqueness and potential of CCR2 PET for AAA patients’ risk-stratification in future clinical settings.

We recently reported the safety of ^64^Cu-DOTA-ECL1i in healthy volunteers and demonstrated the specific detection of CCR2+ cells in patients with pulmonary fibrosis and after myocardial infarction (39, 64). Herein, we expanded these observations and demonstrated the feasibility of ^64^Cu-DOTA-ECL1i imaging patients with AAA who were planned for open repair. Our unique ability to comparatively analyze human PET/CT imaging data and CCR2 content in the surgically harvested AAA tissue provide evidence of tissue validation of the imaging results. In contrast to the age-matched HVs showing low tracer retention in the infrarenal aorta and adjacent vena cava, in AAA patients, ^64^Cu-DOTA-ECL1i demonstrated a heterogeneous and strong uptake along the intraluminal layer of AAA but low signal within the thrombus, suggesting that the majority of tissue inflammation resides within the AAA wall. Particularly, the PET signals were adjacent to the calcified regions on the aortic wall, suggesting binding to atherosclerotic lesions where CCR2+ monocytes and macrophages accumulated (65-67). The substantial expression of CCR2 and its co-localization with CD68 throughout the aneurysmal tissues collected from these patients provided direct support to this contention. The abundant expression of CCR2+ cells in biobanked human AAA and ruptured AAA specimens further highlighted the significance of CCR2 in AAA development and provided unique insights in the likely role of CCR2+ cells in AAA pathogenesis. Although a limited number of RAAA human specimens were assessed, their relatively higher expression of CCR2+ cell comparing to AAA specimens warrants further investigation about the role of CCR2+ cells in the rupture process.

Due to the complex pathophysiology and unknown mechanism of AAA, there are currently no generally accepted medical therapies that directly target and inhibit AAA tissue growth (8, 68, 69). The pathogenic role of proinflammatory monocytes and macrophages makes them attractive targets for AAA therapy (10, 55, 70). Based on the effectiveness of CCR2 inhibition in attenuating immune response in various inflammatory diseases (44, 71-73), we assessed whether CCR2 inhibition could impede AAA expansion and decrease risk of rupture. In contrast to the non-treated MRAAA and FRAAA, RS504393 administration effectively reduced AAA growth and rupture rate in both rat models. Our findings were consistent with the outcomes measured in experimental atherosclerosis models following CCR2 antagonist treatment (44). It is known that macrophages play pathogenic and reparative roles in AAA through their involvement in extracellular matrix remodeling, in promotion and resolution of inflammation, and in various aspects of tissue-healing response. Thus, the timing for CCR2-based anti-inflammatory treatment could be critical in modulating the treatment outcome, which was demonstrated by the survival difference between CCR2i D0 and D3 in MRAAA rats. Since RS504393 inhibits the activation and migration of CCR2+ proinflammatory monocytes from bone marrow, the suboptimal outcome observed in the CCR2i D0 group could likely be explained by the ineffective resolution of initial inflammation (74), highlighting the importance of real-time imaging of macrophage subtypes during anti-inflammatory therapy. Given the sensitivity and specificity of ^64^Cu-DOTA-ECL1i for CCR2 imaging and the reduction of tracer uptake in both RAAA models following CCR2i treatment, our data indicated the feasibility of CCR2 as a theranostic biomarker for AAA management.

There are several limitations in this study. First, all the AAA rat models used were established *via* combined procedures and required unique micro-surgical expertise. A learning curve is needed to achieve consistent and reliable results. Rat AAA pathophysiology was largely driven by the acute, subacute, and chronic phases of inflammation and lacked the contribution of atherosclerotic lesions that are often observed in humans. In contrast to the longitudinal development of AAA in humans, a chronic and clinically relevant animal model that combines both AAA rupture and atherosclerotic disease is needed to better recapitulate human pathophysiology and provide further insights about the theranostic potential of CCR2. Additionally, RS504393 was administered at only one dosage via oral gavage. Optimization of dose administration may improve treatment efficacy in AAA growth and risk of rupture. We also envision that using other CCR2 antagonists with different physiochemical properties (i.e., hydrophobicity) and bioactivities (i.e., affinity, potency) may provide additional impact on the disease process (44, 75). Finally, we acknowledge that there are a limited number of human subjects evaluated that underwent both ^64^Cu-DOTA-ECL1i PET/CT preoperative imaging and then open AAA surgical repair. This unique population is challenging to recruit to clinical trials and our data provide a unique opportunity to integrate PET/CT imaging and *ex vivo* tissue validation to assess the theranostic potential of CCR2 for AAA management.

In conclusion, we identified CCR2 as a unique theranostic biomarker for AAA. We demonstrated that CCR2 content was elevated in AAA tissue in a novel female rat AAA rupture model, an established male rate AAA rupture model, and in human ruptured and non-ruptured AAA tissue. The high specificity of ^64^Cu-DOTA-ECL1i enabled the sensitive detection of CCR2+ cells in these models to assess the risk of AAA rupture. CCR2 inhibitor, RS504393, significantly decreased AAA growth and risk of rupture in both RAAA rat models. The first-in-patient study of ^64^Cu-DOTA-ECL1i PET/CT provided compelling evidence to expand the CCR2 translational research in future research. Collectively, our study provided the impetus to further characterize the role of CCR2 in both diagnosis and treatment of AAAs in human subjects in the future.

## Methods

### Animals

Male and Female Sprague-Dawley rats (200–300g) were obtained from Charles River Laboratories (Wilmington, MA) and used for all described experiments. All rats were housed at 21 °C in a 12/12 hour light/dark cycle and had access to food and water ad libitum. Anesthesia was administered with a mixture of ∼1.5% isoflurane and oxygen for all procedures. The core body temperature was monitored and maintained with a heating pad (37°C). Use of all animal experiments were performed in accordance with relevant guidelines and regulations, and approved by the Institutional Animal Care and Use Committee (IACUC) at Washington University School of Medicine in St. Louis. At the conclusion of studies, live animals were euthanized following IACUC protocols.

### Induction of AAA model and sham-control

Male and female rats were induced to develop infrarenal AAAs via an established model using porcine pancreatic elastase (PPE; 12 U/mL) while one group of female rats were exposed to heat-inactivated PPE (12 U/mL, heated at 90°C for 45 minutes) as surgical control (37). Ventral abdominal wall laparotomy was performed, and the infrarenal abdominal aorta was exposed. A customized polyethylene catheter (Braintree Scientific, Braintree, MA) was introduced through an infrarenal aortotomy, and elastase was infused into the isolated aortic segment for 30 minutes. The exposed aortic segment was dilated to a maximal diameter, and constant pressure was maintained with the use of a syringe pump. Using a video micrometer, the baseline maximum aortic diameter was measured. After 14 days, all rats aortas were re-exposed *via* ventral abdominal laparotomy, maximal aortic diameters were measured, and aortic tissue was harvested for further analysis.

### Promoting AAA rupture in male and female rats

Starting 3 days before elastase exposure and daily thereafter, all male and female rats that were exposed to active PPE also underwent β-aminopropionitrile (BAPN) administration through drinking water (0.3% β-aminopropionitrile in water) to promote AAA rupture. Rats were ≈250 g and drank 25 mL water/day, leading to intake of ≈300 mg BAPN/ (kg·day). Previously, BAPN was shown to effectively promote AAA tissue peak inflammation by day 6, and AAA rupture between days 7 and 14, but unlikely to cause rupture after day 14 if it has not already occurred (76). At the 6 or 14-days’ time points, rats were sacrificed, AAA diameters were evaluated, and aortic tissue was harvested for further analysis. Rats that developed ruptured AAAs (RAAA) during the study period promptly underwent necropsy to confirm and analyze the pathology, whereas those animals that did not rupture by day 14 were identified as the non-ruptured AAAs (NRAAA). Non-operated wild type (WT) and sham-controls female rats used in this study were never exposed to BAPN.

### CCR2 inhibition in male rats

A highly selective CCR2 antagonist, RS504393 (45, 46), was selected to inhibit AAA expansion and rupture, with the aim of testing a preclinical therapy via CCR2 targeted immunomodulation. Male rats that underwent AAA rupture promotion, were divided into three groups based on treatment administration. The first group was composed of a non-treated group of male rats (MAAA; n=25), also considered as treatment controls. A second group of male rats were treated with a CCR2 inhibitor (2 mg/kg)via oral gavage starting at day 0 (PPE exposure) and daily thereafter (CCR2i D0; n = 19). The last group of male rats were treated with CCR2 inhibitor but starting at day 3 post PPE exposure, with an already established AAA (CCR2i D3; n=15) to study the time effect of CCR2 inhibition on aneurysmal development and rupture.

### CCR2 as a target for AAA imaging and therapy in female rats

PET imaging studies were performed in female rats that underwent AAA rupture stimulation (FAAA; n=12) and sham-controls (Sham; n=3) at day 7 and 14 post PPE exposure to follow aneurysm progression. A group of WT female rats (n=3) were also assessed to show radiotracer background retention and non-specific binding.

After establishing the AAA rupture model in females and effectively assessing its CCR2 expressing characteristics, the total of female rats that underwent AAA rupture promotion, including those that were not exposed to PET imaging, were considered as treatment controls (FAAA; n=32). Another group of female rats that also underwent AAA rupture stimulation, were successfully treated with a CCR2 inhibitor via oral gavage, starting at day 3 post PPE exposure (CCR2i; n=15) to also investigate rupture prevention potential of CCR2 inhibition in female rats. *Aortic diameter measurements with ultrasound.* Noninvasive ultrasound (GE, 12 MHz Zonare, Mountain View, CA), was used to evaluate maximal aortic diameter measurements. Relative to baseline aortic diameter prior to PPE exposure, the percentage increase in aortic diameter was evaluated at day 7 and 14 post PPE exposure. As previously described, aortic aneurysms were defined as >100% increase in the aortic maximum diameter relative to baseline diameter (77, 78). *Synthesis and radiolabeling of DOTA-ECL1i.* The CCR2 binding peptide ECL1i (LGTFLKC) was customized using D-form amino acids by CPC Scientific (Sunnyvale, CA). DOTA-ECL1i conjugate was synthesized following our established protocols (37). Copper-64 (^64^Cu, t_1/2_=12.7 hour) was produced by the Washington University Cyclotron Facility. The DOTA-ECL1i conjugate was radiolabeled with ^64^CuCl_2_ (^64^Cu-DOTA-ECL1) following established protocols, and the radiochemical purity (>95%) was determined by radio-HPLC. For human CCR2 imaging, ^64^Cu-DOTA-ECL1 was produced under exploratory investigational new drug application (IND 137620) approved by the US FDA following batch production record in ISO class 7 manufacturing suites under current good manufacturing practices (cGMP) conditions within the biological therapy core facility of Siteman Cancer Center at Washington University. The final product was subject to pre-release quality control analyses before administration to humans. The final product contained 296-366 MBq of ^64^Cu-DOTA-ECL1i with specific activity greater than 18.5 MBq/μg. *Small animal PET/CT imaging and image analysis.* Dynamic PET scan and corresponding CT images were obtained using an Inveon MM PET/CT (Siemens, Malvern, PA) at 45 to 60 minutes after a tail vein injection of ^64^Cu-DOTA-ECL1i (9.5-12.9 MBq per rat) to minimize the effect of blood retention on AAA uptake. PET images were corrected for attenuation, scatter, normalization, and camera dead time, and co-registered with CT images. The PET images were reconstructed with the maximum a posteriori algorithm. The AAA uptake was calculated as standardized uptake value (SUV) in 3 dimensional regions of interest from PET images without correction for partial volume effect using Inveon Research Workplace software (Siemens) (30).

### Histology and immunostaining of rat AAA tissue sections

Aortic tissue was harvested from all animals. AAA tissue was fixed in Histochoice (VWR), and paraffin embedded. Paraffin blocks were sectioned at 5 μm, and deparaffinized. Processing for antigen retrieval was performed with Sodium Citrate solution, pH 6.0, for 10 min. Tissue sections were blocked with 10% serum, and sections were incubated with primary antibody anti-CD68, 1:100 [Bio-Rad, MCA341GA] and CCR2, 1:200 [Novus-Bio, NBP1-48338]. For Immunofluorescence, sections were incubated with donkey anti mouse (Alexa Fluor 647), and donkey anti rabbit (Cy3) [Jackson ImmunoResearch Laboratories]. A Leica THUNDER Imager 3D was used to acquire the images, which were quantified using Image J Software (NIH) and shown as mean intensity. To evaluate AAA tissue morphology and pathology, tissue sections were evaluated using Hematoxylin and Eosin (H&E), Verhoeff-Van Gieson (VVG), and Mason Trichrome (MT) staining using NanoZoomer (Portsmouth, NH) and analyzed blindly in a semi quantitative manner by a clinical pathologist to assess elastin degradation and VSMC loss within the aneurysm wall.

### Autoradiography of rat and human AAA tissues

After PET imaging performed at day 14, groups of FAAA and sham-control female rats were euthanized for specific tissue harvesting and analysis. The thoracic aorta, abdominal aorta and psoas muscle were collected and infused with saline. The aortic tissue was then sliced longitudinally and fixed in paraformaldehyde for 2 hours, followed by 2.5-minute rinses with 0.9% sodium chloride. All tissues were placed on a charged phosphor screen and exposed overnight. For human AAA tissue, de-identified paraffin-embedded specimens were acquired from the Vascular Surgery Department Bio Bank under the approval from the Human Research Protection Office of Washington University. All slides were first deparaffinized and rehydrated through a series of graded alcohols. Tissues were boiled in citrate-based Unmasking Solution (pH 6; Vector Laboratories) for antigen retrieval and then blocked with 1% BSA and 0.1% Tween in 1X PBS to minimize nonspecific binding. The slides were immersed in a Coplin jar containing ^64^Cu-DOTA-ECL1i (approximately 1.0 MBq/mL) in MilliQ water and incubated for 15 minutes with shaking. The radiotracer was decanted, and the slides were rinsed with MilliQ water and exposed overnight on a charged phosphor screen. All autoradiography assays were performed with a GE Typhoon FLA 9500 Variable Mode Laser Scanner at 50-micron resolution. ImageJ software was used for the quantification of ^64^Cu-DOTA-ECL1i uptake in AAA tissue samples. A region of interest was drawn around the entire tissue section. Mean pixel intensity was compared between AAA samples and sham-controls.

### Human studies

Human ^64^Cu-DOTA-ECL1i PET/CT study was approved by the Washington University School of Medicine Institutional Review Board (IRB, 202008190). Informed consent was obtained from all subjects prior to enrollment. Four AAA patients between the age of 56-70 years old scheduled for open repair were recruited. The ^64^Cu-DOTA-ECL1i PET/CT scans were carried out within 7-14 days of their scheduled surgery to minimize changes in CCR2 expression measured at the time of AAA tissues collection. CT angiography (CTA) with contrast agent were also performed. Four healthy volunteers between 32 and 75 years old with documented absence of AAA by screening ultrasound that was previously obtained as part of standard care were also recruited as controls. Patient exclusion criteria includes inability to receive and sign informed consent, with Stage ≥4 chronic renal failure (calculated by modification of diet in renal disease eGFR equation [to minimize confounding imaging variables]), documented allergy to iodinated contrast and/or shellfish, with an unstable clinical condition that in the opinion of the principal investigators or designee precludes participation in the study, inability to tolerate 60 minutes in a supine position with arms down at sides, as necessary for PET/CT, positive pregnancy test or lactating, symptomatic/recently treated coronary disease, cancer requiring oncologic management, autoimmune/inflammatory diseases (e.g., rheumatoid arthritis or multiple sclerosis) that are known to have increased associated CCR2 expression, no known aortoiliac occlusive disease.

### Imaging protocol

All subjects underwent a 60-minute dynamic PET/CT scan (Biograph Vision 600, Siemens Healthcare, Knoxville, TN, USA) with the field-of-view set over the abdominal aorta following the intravenous injection of 296-366 MBq of ^64^Cu-DOTA-ECL1i in 10 mL of saline. Each PET scan was accompanied by a low-dose CT scan for PET attenuation correction. A CT angiogram (CTA) was acquired if the patient standard of care CTA is older than 2 weeks. Blood samples were collected at 5min and 60 min post radiotracer injection to analyze metabolites (39). *Kinetic modeling*. We performed a kinetic analysis of ^64^Cu-DOTA-ECL1i PET scans using multiple image-derived input functions (IDIFs) obtained from aorta above AAA. Since AAA patients might have systemic inflammation within the aorta, we also selected upper vena cava to generate IDIF. Time-activity curves (TACs) were extracted from 29 sequential PET frames of increasing duration within the 0-60 minute dynamic scan, focusing on both the blood input functions and the AAA lesion. The distribution volume (DV) was calculated using Logan plot and IDIF data as representative of the blood input functions using PMOD version 4.1. The Logan plot technique utilizes a compartmental model and linear regression to examine the pharmacokinetics of tracers with reversible uptake (eq.1). Particularly, DV values pertains to the apparent volume within the body where a drug is distributed compared to its concentration in the plasma. In our context, a value close to 1 indicates that the tracer’s target volume aligns closely with that of the blood. All IDIFs were corrected for metabolite effects. For HV subjects, regions of interest were draw at abdominal aorta above the bifurcation and adjacent vena cava.

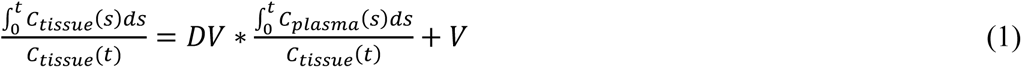

### ELISA and cytokine array

AAA tissue protein was extracted using RIPA buffer with proteinase inhibitor (Sigma #MCL1). Protein quantification was done by Bradford assay. For each AAA tissue samples 25ug of protein was analyzed for CD68 (MyBioSource, MBS705029), CCR2 (antibodies-online, ABIN6974763) and Cytokine multiplex assay (Millipore, RECYTMAG-65K) using manufacturer instructions.

### Statistics

All data are presented as the mean ± SD. Multiple group comparisons were performed using One-way ANOVA with a Tukey test. Individual group differences were determined using a 2-tailed Mann-Whitney test. Data was considered statistically significant with p < 0.05. Kaplan-Meier curve was generated to assess the survival of BAPN-exposed animals. GraphPad Prism 9 (La Jolla, CA) was used for all statistical analyses and graphical data representations.

### Study approval

Animal experiments were conducted in accordance with approved IACUC protocols (Washington University). Human biobanked AAA tissues were obtained in accordance with an approved IRB protocol (201309043).

## Data Availability

All data produced in the present study are available upon reasonable request to the authors

## Author contributions

SEB, SSD, LD, BA, GSH, DS, HL, XZ, XG, KH, DT, LM, and CL conducted experiments. SEB, CC, CL, JZ, JI, RL, RJG, SJE, MAZ, and YL contributed to study design, reviewed and revised the manuscript. SEB, CL, YK, MAZ, and YL analyzed and interpreted the data. SEB, MAZ, and YL wrote the manuscript. RJG, SJE, MAZ, and YL designed and supervised the study.

## Acknowledgments

We would like to thank the preclinical imaging facility for animal PET/CT scans, which is supported by NIH/NCI Siteman Cancer Center (SCC) Support Grant P30CA091842, NIH instrumentation grants S10OD018515 and S10OD030403, and internal funds provided by Mallinckrodt Institute of Radiology. The human imaging was carried out in the Center for Clinical Imaging Research supported by Mallinckrodt Institute of Radiology. RJG is supported by grants from the NIH (R01HL153436, R01HL150891, P41EB025815). MAZ is supported by grants from the NIH (R01HL153436, R01HL150891, R01HL159803, R0153262). YL is supported by grants from the NIH (R01HL153436, R01HL150891, P41EB025815, R35 HL145212).

Address correspondence to: Yongjian Liu, Mallinckrodt Institute of Radiology, 510 S. Kingshighway Blvd, Campus Box 8225, Washington University in St. Louis, St. Louis, Missouri 63110, USA. Or to: Mohamed Zayed, 4921 Parkview Place, Campus Box 8109, Washington University in St. Louis, St. Louis, Missouri 63110, USA.

