## supplemental material for "Chemokine Receptor 2 Is A Theranostic Biomarker for Abdominal Aortic Aneurysms"

**This PDF file includes:**

Supplemental Figures 1-7

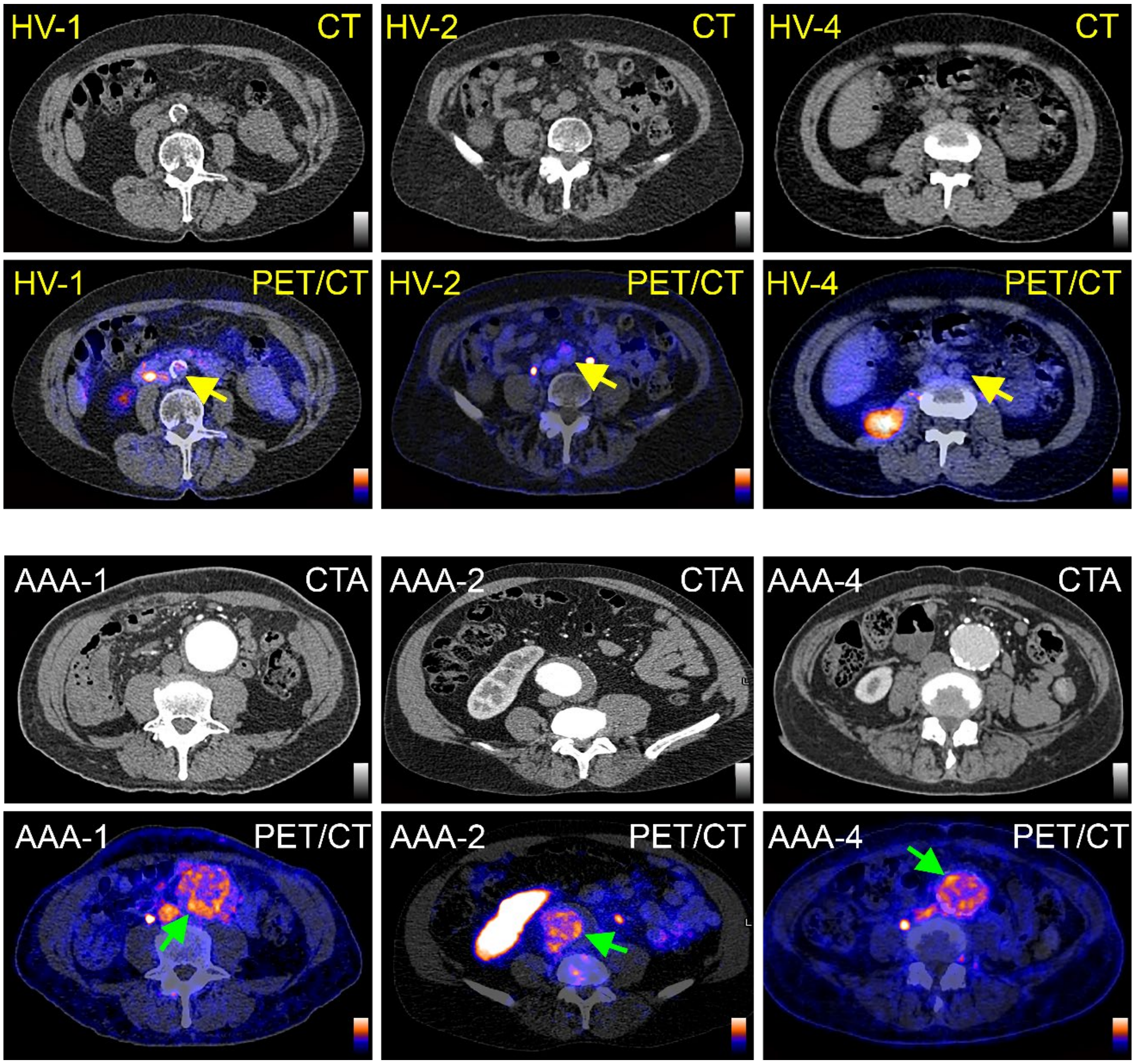


**Supplemental Figure 1. $^{64}$Cu-DOTA-ECL1i PET/CT images in healthy volunteers (HVs) and patients with AAA.** Yellow arrow: transverse view of infrarenal aortas in HVs. Green arrow: transverse view of AAAs.

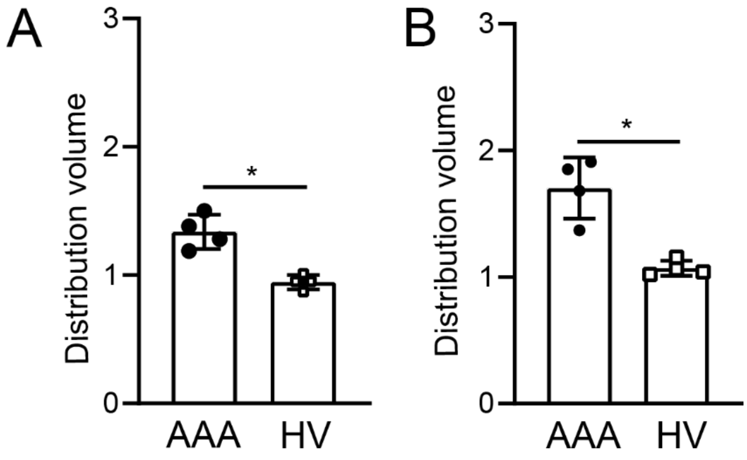


**Supplemental Figure 2. Distribution volume values for AAA patients and HV subjects.** (**A**) Multiple image-derived input functions obtained from upper aorta. (**B**) Multiple image-derived input functions obtained from upper vena cava.

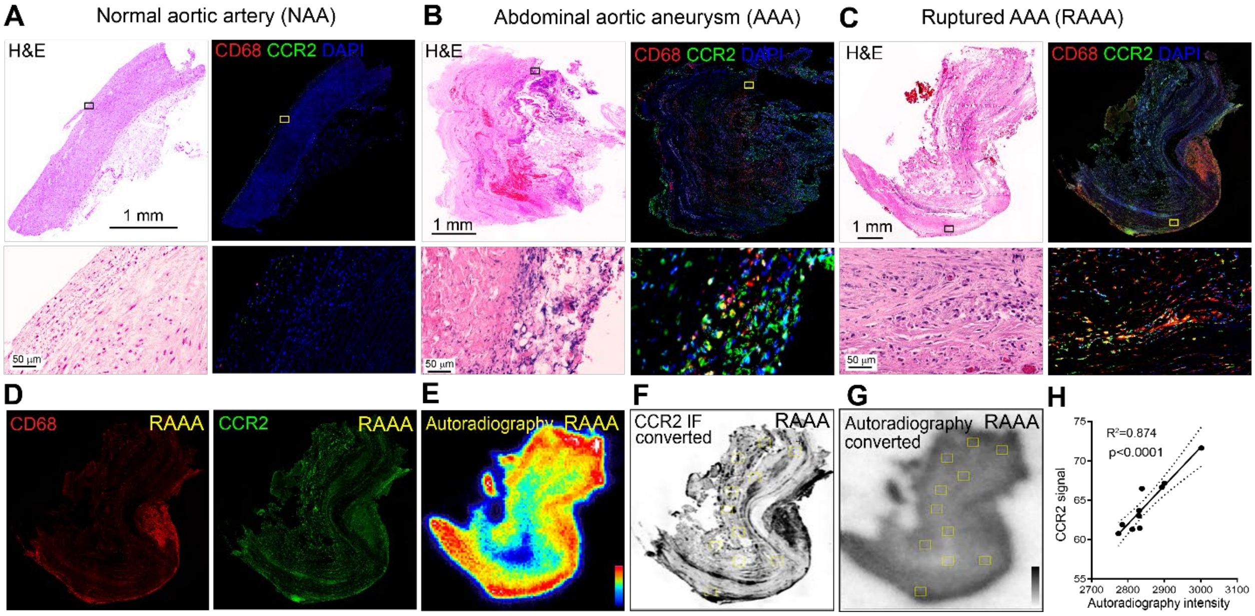


**Supplemental Figure 3. Characterization of CCR2 expression in human aortic artery tissues.** (**A**) Picture showing a human tissue of a healthy abdominal aortic artery (AA) stained with H&E and IF with CD68 and CCR2 specific antibodies. (**B**) Human sample of a non-ruptured abdominal aortic aneurysm (AAA) stained with H&E and IF with CD68 and CCR2 specific antibodies. (**C**) Human sample of a recently ruptured abdominal aortic aneurysm (RAAA) harvested during open surgical repair intervention, stained with H&E and IF with CD68 and CCR2 specific antibodies. (**D**) Representative picture of CD68+ and CCR2 + cells wall infiltration of RAAA human tissue respectively. (**E**) $^{64}$Cu-DOTA-ECL1i autoradiography of RAAA tissue. Converted low-resolution images of IF staining of CCR2 (**F**) and autoradiography (**G**). The two sets of images were co-registered with representative regions-of-interest (ROIs, yellow squares). (**H**) Signals extracted from the ROIs of the two sets of images showed linear correlation.

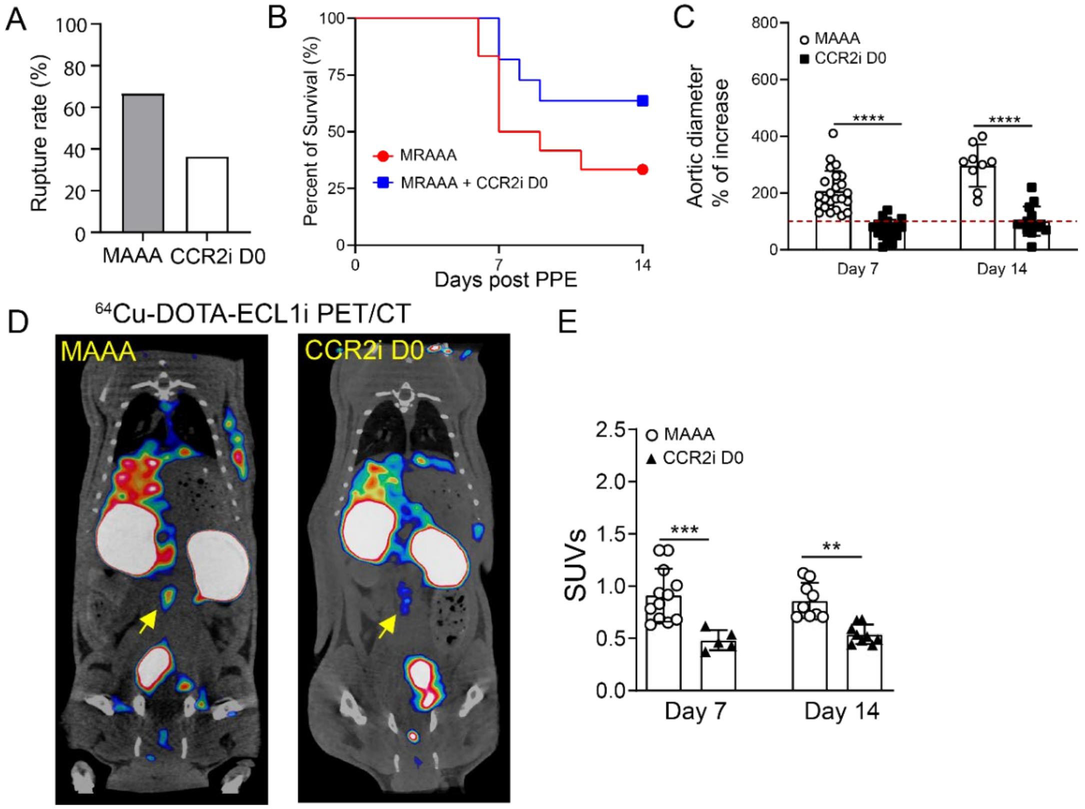


**Supplemental Figure 4. Assessment of CCR2 inhibition starting at day 0 on AAA progression and rupture in a MAAA rat model.** (**A**) Comparison of AAA rupture rates between MAAA rats without treatment and those treated with CCR2i starting at day 0 (CCR2i D0). (**B**) Kaplan-Meier curve demonstrating significantly improved survival of MAAA rats following CCR2i D0 treatment. (**C**) Percent aortic diameter increase in MAAA vs CCR2i D0 groups at day 7 and day 14, respectively. Red dotted line is set at 100%. (**D**) Representative $^{64}$Cu-DOTA-ECL1i PET/CT images at day 14 post PPE exposure showed specific and intense detection of AAA (yellow arrow) in MAAA, compared with the low trace accumulation in the CCR2i D0 treated group. (**E**) Quantitative tracer uptake in MAAA rats without and with CCR2i D0 treatment at day 7 and day 14 post PPE. Data presented as Mean ± SD. *$p < 0.05$, **$p < 0.01$, ***$p < 0.001$, **** $p<0.0001$.

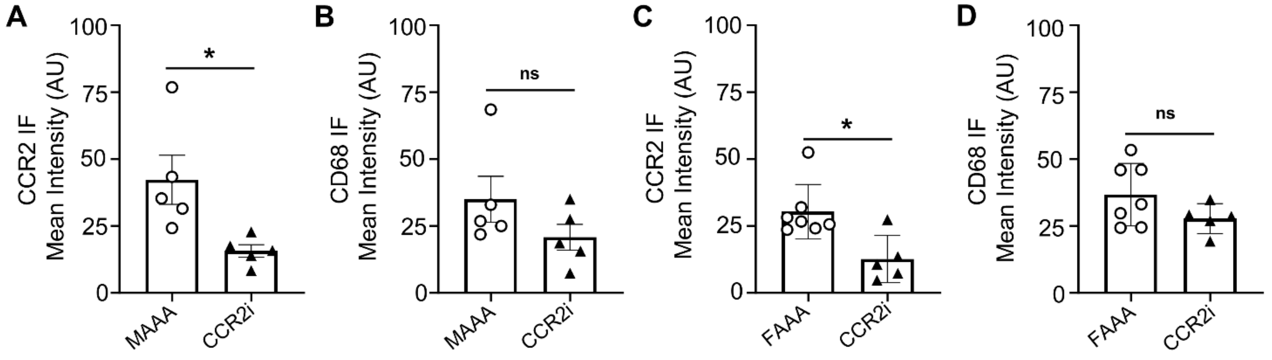


**Supplemental Figure 5. Quantification of immunofluorescence intensity of CCR2+ cells and CD68+ macrophages within AAA wall following CCR2 inhibition in MAAA and FAAA models.** (**A**) CCR2+ cells in non-treated MAAA vs CCR2i D3 (42±20 vs 16±5; respectively) in male rats. (**B**) CD68+ macrophages in non-treated MAAA vs CCR2i D3 (35±19, n=5 vs 21±10) in male rats. (**C**) CCR2+ cells in non-treated FAAA vs CCR2i D3 (30±10 vs 13±9) in FAAA. (**D**) CD68+ macrophages in non-treated FAAA vs CCR2i D3 (37±12, n=7 vs 28±6) in FAAA. Data presented as Mean ± SD. *p < 0.05, **p < 0.01, ***p < 0.001, **** p<0.0001.

.

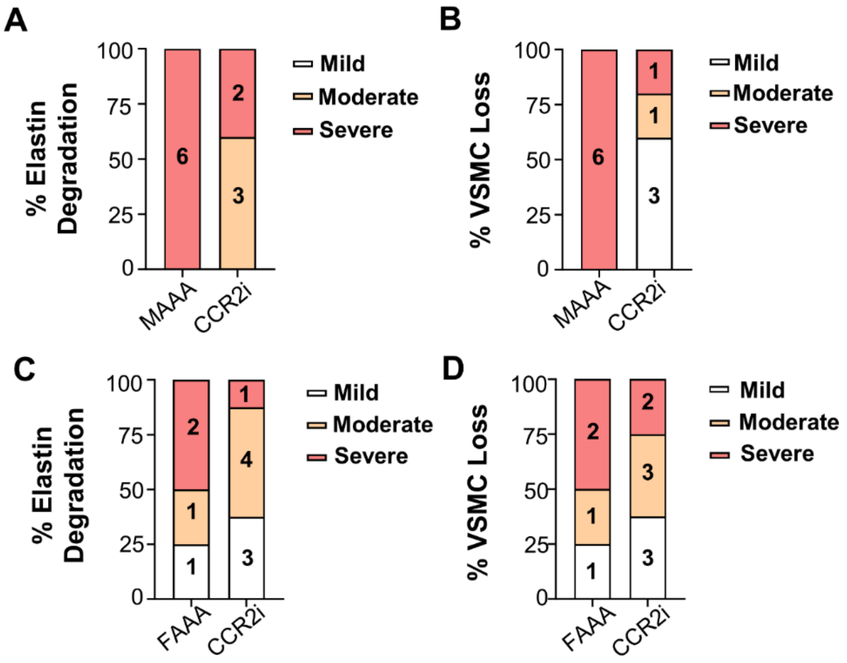


**Supplemental Figure 6. Histopathological characterization of AAA in male and female rats following CCR2i.** (**A**) Elastin degradation assessment in non-treated MAAA (n = 6 for severe) vs CCR2i in MAAA model (n= 3 for moderate and n = 2 for severe). (**B**) VSMC loss assessment in non-treated MAAA (n=6 severe) vs CCR2i (n=3 mild, n=1 moderate and n = 1 severe) in MAAA model. (**C**) Elastin degradation assessment in non-treated FAAA (n=1 mild, n= 1 moderate and n=2 for severe) vs CCR2i in FAAA model (n= 3 mild, n=4 moderate and n=1 severe). (**D**) VSMC loss assessment in non-treated FAAA (n=1 mild, n=1 moderate, n=2 severe) vs CCR2i in FAAA (n=3 mild, n=3 moderate and n = 2 severe).

A

| | Elastin Degradation | VSMC loss |
|---|---|---|
| **Mild** | Mostly intact, with less than 10% disruption | Decrease nucleus density less than 10% |
| **Moderate** | Shredded elastin layers, 10-70% disruption | 10-50% |
| **Severe** | More than 70% disruption | More than 50% |

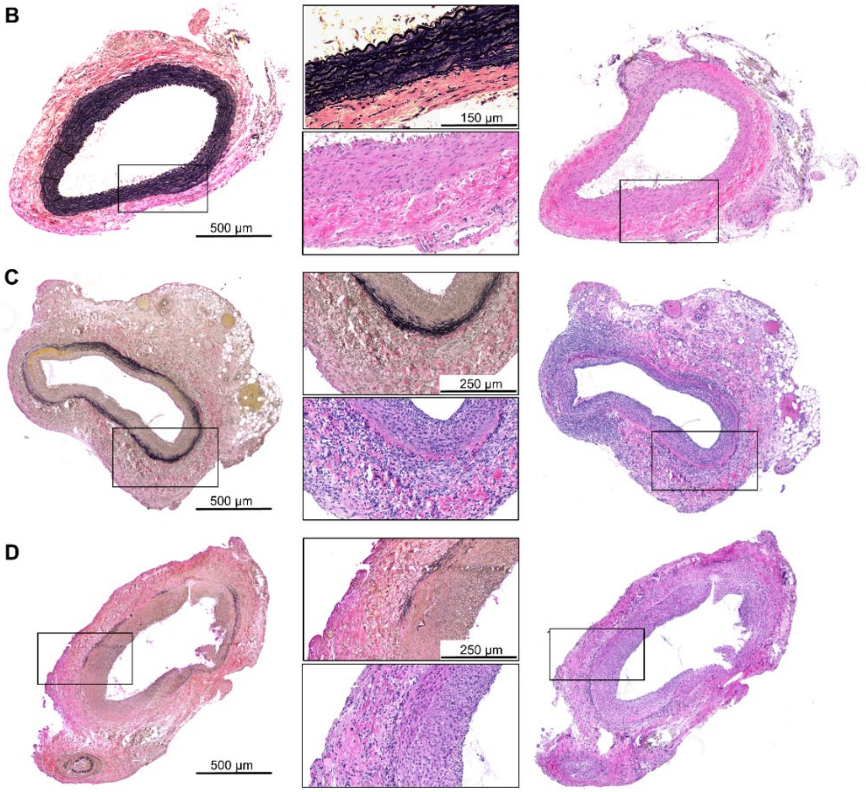


**Supplemental Figure 7. Histopathological grading system for rat AAA tissue.** (A) Table with grading system and measured parameters: Elastin degradation (mild = mostly intact elastin fibers with less than 10% of disruption; moderate = shredded elastin layers with 10-70% of disruption and severe more than 70% of elastin fiber disruption) and VSMC loss (mild = decreased nucleus density; moderate = 10-50% of decrease in nucleus density; and severe >50% decrease in nucleus density, VSMC mostly gone). VVG and H&E staining of abdominal aortas (cross-sectional) with 5x magnification and 10x magnification to visualize elastin degradation and VSMC loss in rats aortic tissue with either (B) mild (C) moderate and (D) severe example of analysis.
